# SARS-CoV-2 serostatus of healthcare worker in the Austrian state Vorarlberg between June 2020 and January 2021

**DOI:** 10.1101/2021.02.19.21252045

**Authors:** Michele Atzl, Axel Muendlein, Thomas Winder, Peter Fraunberger, Eva-Maria Brandtner, Kathrin Geiger, Miriam Klausberger, Mark Duerkop, Lukas Sprenger, Beatrix Mutschlechner, Andreas Volgger, Magdalena Benda, Luciano Severgnini, Johannes B. Jaeger, Heinz Drexel, Alois Lang, Andreas Leiherer

**Author notes:** **Address for correspondence** Andreas Leiherer, Vorarlberg Institute for Vascular Investigation and Treatment (VIVIT), Academic Teaching Hospital Feldkirch, Feldkirch, Austria, Carinagasse 47, A-6800 Feldkirch, Austria. **Funding and disclosures** This work received a particular funding from FFG (project number 880956). No potential conflicts of interest relevant to this article were reported by M.A., A.M., T.W., P.F., E.M.B., K.G., M.K., M.D., L.Sp., B.M., A.V., M.B., L.Se., J.J., H.D., A.La., and A.Le. **Data sharing statement** The data that support the findings of this study are available from the corresponding author upon reasonable request.

## Abstract

**Background:** Austria, and particularly its westernmost federal state Vorarlberg, developed an extremely high COVID-19 incidence rate in November 2020. Health care workers (HCW) may be at higher risk of contracting the disease within the working environment and therefore the seroprevalence in this population is of particular interest. Here, we analyzed SARS-CoV-2-specific antibody response in Vorarlberg HCW in a prospective cohort study.

**Methods:** A total of 395 HCW have been tested at three different time points for the prevalence of anti-SARS-CoV-2 IgG antibodies specific for NP and RBD. Enrollment started in June 2020 (t_1_), two months after the end of the first wave. Re-testing took place between October to November at the beginning of the second wave (t_2_), and again at the end of the second wave in January 2021 (t_3_).

**Results:** At t_1_, 3% of HCW showed a strong IgG-specific responses to either NP or RBD. At t_2_, the rate increased to 4%, and after the second wave in January 2021, 14% had a strong response, which was assessed to be stable for up to ten months. The amount of HCW with anti-SARS-CoV-2 IgG antibodies was 38% higher than the number of infections found by RT-PCR.

**Conclusion:** We found low numbers of SARS-CoV-2-seropositive HCW in a hotspot setting after the first wave but a very high increase during the second massive wave. Though the seroprevalence in HCW was comparable to the general population. Our findings offer support for the routine application of serological testing in management of the ongoing COVID-19 pandemic.

**Main summary:** A relatively low percentage of 3% SARS-CoV-2 seropositive HCW with strong IgG-specific antibody responses was found in the Austrian federal state Vorarlberg after the first wave increasing to 14% after the second massive wave lasting until January 2021.

## Introduction

In March 2020 the coronavirus disease 2019 (COVID-19) was declared a global pandemic by the World Health Organization (WHO), with Europe at the time as the epicenter. The high numbers of cases and associated mortality first overwhelmed health care services in Northern parts of Italy (1). Several independent introducing events, mainly from Northern Italy have most likely contributed to clusters in Austria (2) and further enforced the spread in many other European countries (3) during the so called first wave in March 2020. During the second and by far higher wave, peaking in Austria in November, Austria developed the highest incidence rate worldwide (4) and the federal state of Vorarlberg, despite a low degree of urbanization, reported one of the highest rates in Austria (5).

Health care workers (HCW) are on the first line of defense and have a high risk of becoming infected and infecting others with SARS-CoV-2 (6). This has been first demonstrated in China (7) and has been confirmed in early reports from Italy, where HCW have represented 9% of total cases (1).

In contrast to real time reverse transcription polymerase chain reaction (RT-PCR) assays detecting SARS-CoV-2 for only 2-3 weeks after infection (8), the immunoglobulin (Ig) G-specific response to SARS-CoV-2 epitopes is typically detectable in serum about two weeks after symptom onset (9). At least 95% of PCR-confirmed SARS-CoV-2 infected patients develop specific anti-SARS-CoV-2 antibodies (10). The receptor binding domain (RBD) of the spike protein has meanwhile become the most common antigen of seroconversion assays, as it has received FDA emergency approval (11) and has also been shown to correlate well with neutralizing activity (10, 12–14).

This study thus investigates the dynamics of IgG-specific response against RBD and the nucleocapsid protein (NP) of SARS-CoV-2 in serial serum samples collected from 395 HCW after the first wave (June – August 2020), at the beginning of the second massive wave (October 2020), and the end of the second wave (January 2021) using enzyme linked immunosorbent assay (ELISA).

## Methods

### Study subjects

This study comprises 395 participants of mainly Caucasian origin with a median age of 42 (min. 18 – max. 64) years working as HCW in Vorarlberg, the westernmost federal state of Austria. All participants are employed by either of the Vorarlberg state hospitals and 174 (44%) at a COVID-19-specialized hospital.

Study enrolment was voluntary and at no charge for the participants. All subjects reported to be in healthy condition. Inclusion rested solely towards the purpose of antibody determination and was not influenced or motivated by the presence of signs or symptoms of disease. At the time point of recruiting, participants completed a survey form which prompted for demographic information as well symptoms of COVID-19 infection in the three months prior to collection of serum sample. Additionally, data on SARS-CoV-2-specific RT-PCR test were collected, which had been ordered by the hospital at any suspicion of a possible infection. All participants gave written informed consent and the present study conforms to the ethical guidelines of the 1975 Declaration of Helsinki and has been approved by the Ethics Committee of Vorarlberg (EK-2-4/2020).

After the first wave in March 2020 and after the first harsh lockdown in Austria (16^th^ of March to 30^th^ of April) blood samples were collected. Collection took place between 26^th^ of June and 19^th^ of August 2020 and is referred to as time point 1 (t_1_). Identical criteria were applied for the second round of sampling between 2^nd^ October and 13^th^ November (t_2_) and the third round between 7^th^ and 20^th^ January 2021 (t_3_). Thus, sampling at t_2_ took place mostly at the beginning of the second wave 2020 and at t_3_ after the second wave, during the third harsh lockdown in Austria (17^th^ November to 6^th^ December). A summary of the study timeline is given in **figure 1**. Data on 7-day incidence were obtained from open government data by the Austrian Federal Ministry of Social Affairs, Health, Care and Consumer Protection (15). Only 5 out of 395 participants were missing at t_2_ and 24 at t_3_ due to end of employment, withdrawal of consent, or due to other reasons. Hence, the follow-up rate at t_2_ was 99 % and 94% at t_3_.

**Figure 1.**
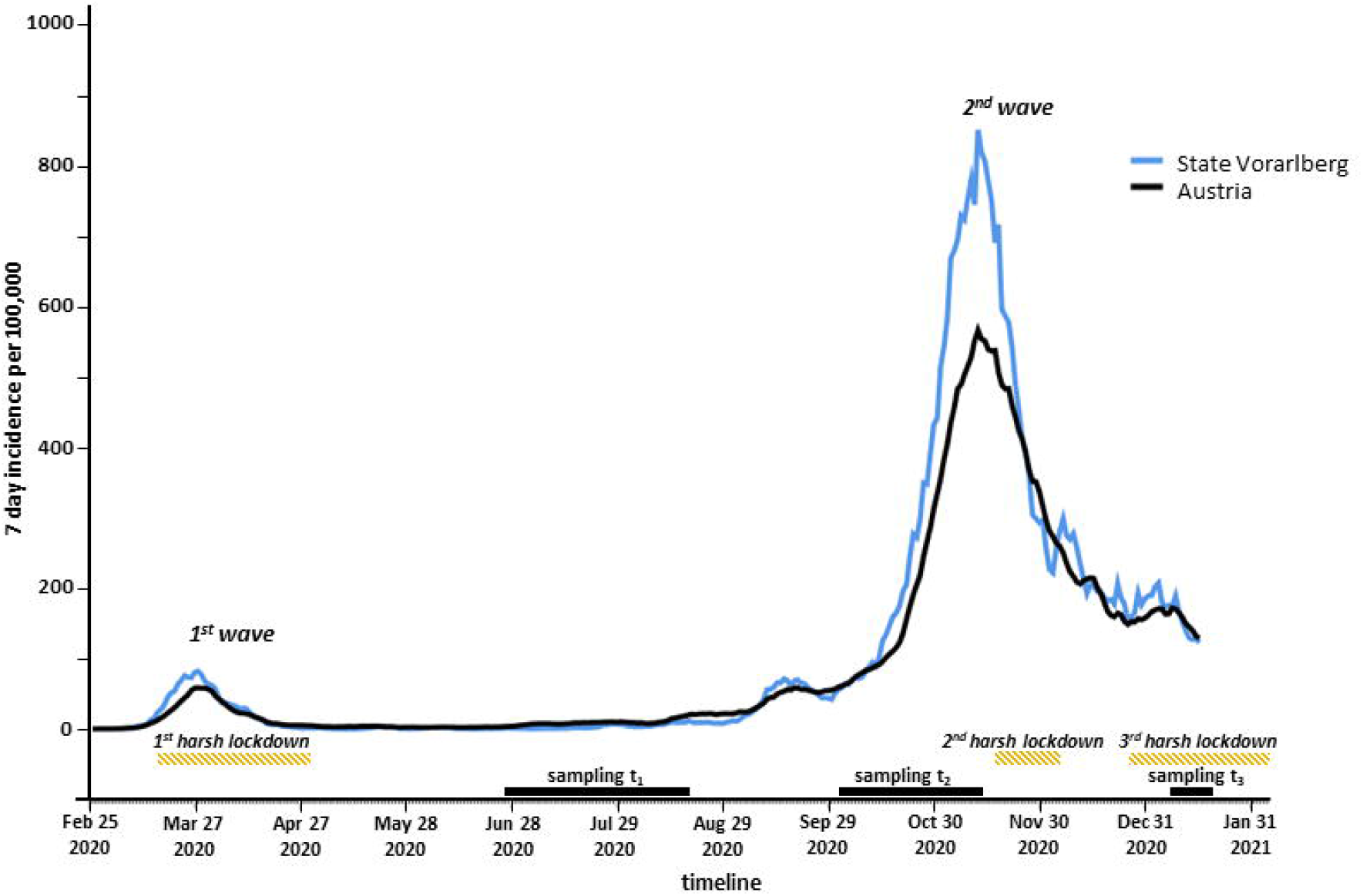
Study timeline. The figure presents the 7-day incidence per 100,000 inhabitants in Austria and in the federal state of Vorarlberg between February 2020 and January 2021. The time points of sampling (t_1_, t_2_, and t_3_; solid black line) and of harsh lockdown (hatched line) are marked.

### Study data and laboratory analyses

Study data were collected and managed using REDCap electronic data capture tools (16, 17) hosted at VIVIT. Acute SARS-CoV-2 infection was determined by virus detection through RT-PCR of nasopharyngeal swabs at the Institute of Pathology, Academic Teaching Hospital Feldkirch (Feldkirch, Austria). At each time point, venous blood was collected, processed, and anti-SARS-CoV-2 antibodies were detected in human serum via an ELISA specifically detecting IgGs directed against the recombinant NP RBD (5600100 Technozym, Technoclone, Vienna, Austria, (13)) according to the manufacturer’s protocol. Concentrations were calculated according to internal calibration standards using the Xlfit software package (Version 5.3.1.3, IDBS) with 1 U/mL representing 100 ng/ml of a SARS-specific antibody (18).

According to manufacturer’s protocol, values <5 U/mL were referred to as normal or background range representing no SARS-CoV-2-specific antibody response. Values ≥5 U/mL were referred to as positive responses. The 5 U/mL cutoff was defined on basis of criteria suggested by the Youden index and the 99^th^ percentile method (19). Values ≥5 and <9 U/mL for anti-SARS-CoV-2 RBD-specific antibody response or ≥5 and <8 U/mL for anti-SARS-CoV-2 NP-specific antibody responses were referred to as a moderate positive response. In terms of the prevalence nature of the study, a higher cut-off of ≥9 U/mL was chosen for anti-SARS-CoV-2 RBD IgG and ≥8 U/mL for anti-SARS-CoV-2 NP IgG to increase specificity, as proposed by the manufacturer and by a previous study (19). Values ≥9 and ≥8 U/mL, respectively were thus referred to as a strong positive response. IgG concentration was measured at time points t_1_, t_2_, and t_3_. Participants whose antibody levels increased between time points from background to moderate, from moderate to strong, or from background to strong response were referred to as converters. Participants with (i) a moderate or strong response at an earlier time point and (ii) no conversion during following time points and (iii) a declined or unchanged response (including also marginally increased responses not higher than 10% or 1 U/mL, respectively) were referred to as moderate or strong response decliner, respectively. The half-life of antibody response as well as the time until antibody response has dropped under the 5 U/mL threshold for seropositivity was extrapolated, assuming an exponential decline.

### Statistical analysis

Differences in baseline characteristics were tested for statistical significance with Chi-squared tests for categorical variables, with Mann-Whitney-U tests for continuous, not normally distributed, and unpaired continuous variables, and with Wilcoxon tests for continuous, not normally distributed, and paired variables, respectively. Correlation analyses were performed calculating nonparametric Spearman rank correlation coefficients. Results are given as mean if not denoted otherwise, and p-values of 0.05 were considered significant. All statistical analyses were performed with SPSS 26.0 for Windows (IBM corp., USA), and R statistical software v. 3.5.1 (http://www.r-project.org).

## Results

### Seroprevalence between June 2020 and January 2021

The study timeline is summarized in **figure 1** and the characteristics of our study participants in **table 1** (further data on residence and profession are summarized in **supplemental table 1**). The anti-SARS-CoV-2 specific IgGs against RBD and NP were assessed at three time points, after first wave (t_1_), at the beginning of second wave (t_2_), and after second wave (t_3_). The respective mean concentrations, the correlation of RBD- to NP-specific IgGs, as well as the proportion of seropositive subjects (5 U/mL cut-off) and in particular the seropositive subjects with a strong response (strong responder: 9 U/mL cut-off) are summarized in **table 2 and figure 2** for the three time points t_1_, t_2_, and t_3_. Overall, 73 (18%) out of all 395 HCW have been tested at least once positive at any time point (t_1_, t_2_, or t_3_) during the study.

**Table 1.**
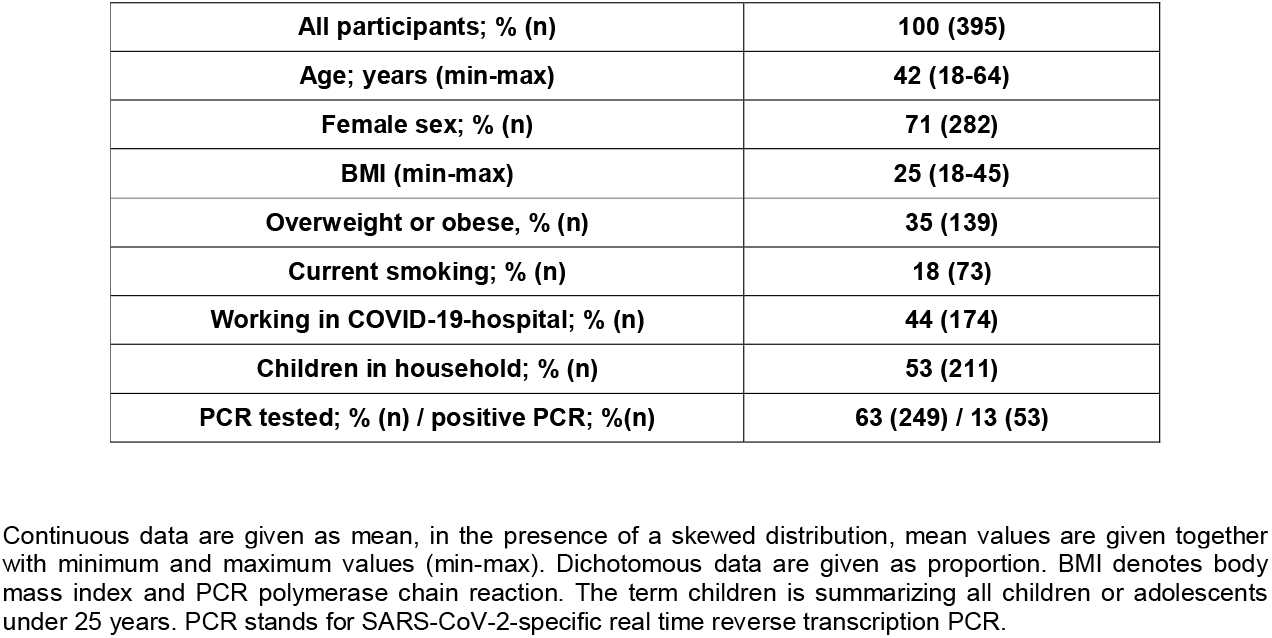
Characteristics

Characteristics
|  |  |
| --- | --- |
| **All participants; % (n)** | **100 (395)** |
| **Age; years (min-max)** | **42 (18-64)** |
| **Female sex; % (n)** | **71 (282)** |
| **BMI (min-max)** | **25 (18-45)** |
| **Overweight or obese, % (n)** | **35 (139)** |
| **Current smoking; % (n)** | **18 (73)** |
| **Working in COVID-19-hospital; % (n)** | **44 (174)** |
| **Children in household; % (n)** | **53 (211)** |
| **PCR tested; % (n) / positive PCR; %(n)** | **63 (249) / 13 (53)** |

Continuous data are given as mean, in the presence of a skewed distribution, mean values are given together with minimum and maximum values (min-max). Dichotomous data are given as proportion. BMI denotes body mass index and PCR polymerase chain reaction. The term children is summarizing all children or adolescents under 25 years. PCR stands for SARS-CoV-2-specific real time reverse transcription PCR.

**Table 2.** Antibody response during study

|  | participants |  | RBD | NP | RBD-NP correlation |
| --- | --- | --- | --- | --- | --- |
| **t<sub>1</sub>** | **all HCW** | 100%<br>(n=395) | 1.66<br>(0.12-0.89) U/mL | 1.40<br>(0.15-0.98) U/mL | r=0.243<br>p<0.001 |
|  | **seropositive HCW** | 6%<br>(n=24) | 18.24<br>(1.55-10.54) U/mL | 13.45<br>(1.94-22.71) U/mL | r=0.270<br>p=0.201 |
|  | **seropositive HCW with strong response** | 3%<br>(n=12) | 32.29<br>(5.00-35.25) U/mL | 24.23<br>(9.35-35.35) U/mL | r=-0.028<br>p=0.931 |
| **t<sub>2</sub>** | **all HCW** | 100%<br>n=390 | 2.78<br>(0.04-0.84) U/mL | 1.59<br>(0.00-0.86) U/mL | r=0.305<br>p<0.001 |
|  | **seropositive HCW** | 6%<br>(n=25) | 35.55<br>(4.68-57.16) U/mL | 17.04<br>(2.10-25.30) U/mL | r=0.338<br>p=0.098 |
|  | **seropositive HCW with strong response** | 4%<br>(n=16) | 52.38<br>(7.51-114.10) U/mL | 24.98<br>(5.71-39.98) U/mL | r=0.206<br>p=0.444 |
| **t<sub>3</sub>** | **all HCW** | 100%<br>(n=371) | 5.17<br>(0.10-1.09) U/mL | 4.52<br>(0.22-1.50) U/mL | r=0.474<br>p<0.001 |
|  | **seropositive HCW** | 17%<br>(n=62) | 28.69<br>(6.57-33.54) U/mL | 23.60<br>(4.93-23.59) U/mL | r=0.448<br>p<0.001 |
|  | **seropositive HCW with strong response** | 14%<br>(n=52) | 33.20<br>(10.39-45.08) U/mL | 27.57<br>(7.71-28.30) U/mL | r=0.347<br>p=0.012 |

The table summarizes the concentration of SARS-CoV-2 receptor binding domain (RBD) - and nucleocapsid protein (NP) - specific antibody response at the respective time point given as mean (with interquartile range). Correlation (r) is given together with the p-value according to spearman test. Seropositive HCW (comprising a moderate and a strong response) had a concentration of $\geq 5$ U/mL for either RBD or NP-response. Seropositive with a strong response were characterized by a concentration of either $\geq 9$ U/mL for RBD or $\geq 8$ U/mL for NP.

**Figure 2.**
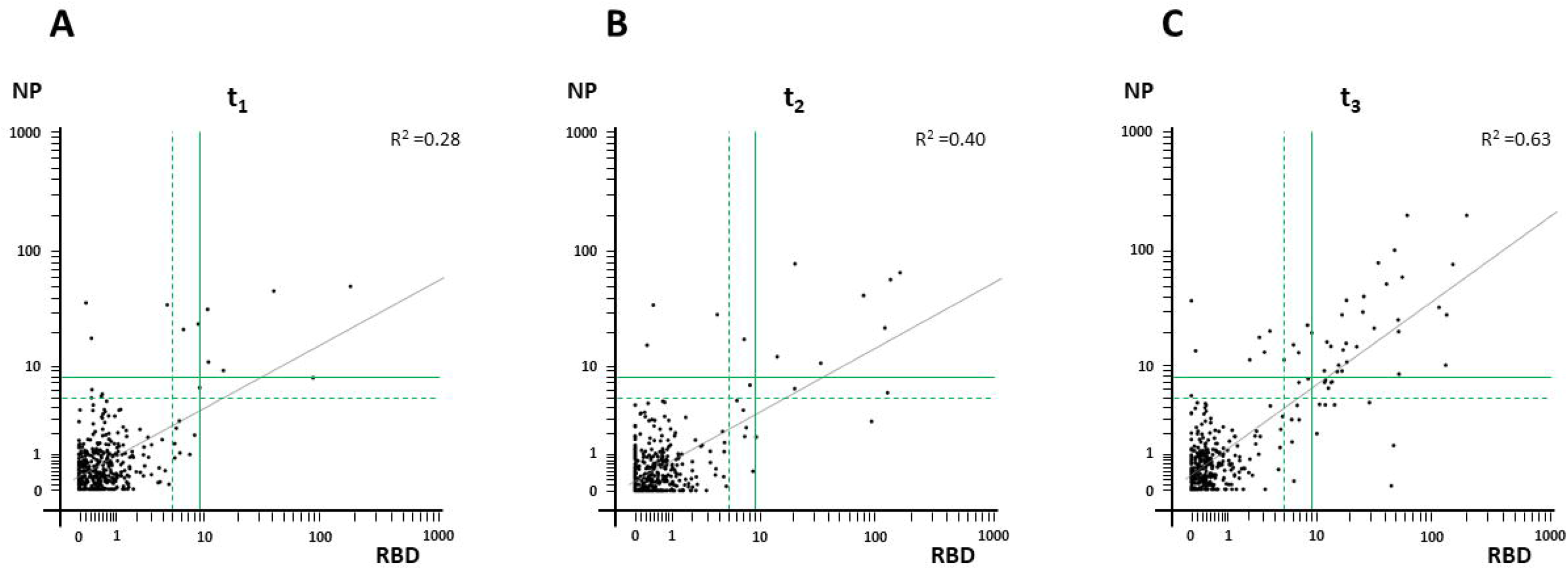
Concentration and spread of RBD- and NP-specific IgG response. SARS-CoV-2-specific anti-RBD and anti-NP-specific IgG response of study participants is depicted at study time point t_1_ (A), t_2_ (B), and t_3_ (C). A reference range of 0-5 U/mL representing no response is separated from a moderate positive response (≥5 and <9 U/mL for anti-RBD IgG and ≥5 and <8 U/mL for anti-NP IgG) by a dashed green line and from a strong positive response (≥ 9 U/mL for anti-RBD and ≥ 8 U/mL for anti-NP) by a solid green line. The solid grey line represents a linear regression line (R^2^).

### Change of antibody response during study

The range of RBD- and NP-specific antibody response between time point t_1_ and t_3_ is depicted in **figure 3** and the change is summarized in **supplemental table 2**. Overall, the RBD- and NP-specific IgG concentration increased during the study. Between t_1_ and t_3_, 44 (12%) seroconverted to a strong response (t_1_-t_3_-strong response converter) and 6 (2%) to a moderate response (t_1_-t_3_-moderate response converter). Out of these 44 t_1_-t_3_-strong response converter, 43 converted from no response at t_1_ to a strong response at t_3_, and only one participant from a moderate response to a strong response. The mean increase of these 44 t_1_-t_3_-strong response converter was 42.3-fold for RBD- and a 43.7-fold for NP-specific antibody response; for the 6 t_1_-t_3_-moderate converters 3.5-fold and 2.3-fold, respectively. Further, 19 HCW were found to have a declined antibody response between t_1_ and t_3_ (t_1_-t_3_- decliner). Of these, nine had a strong response at t_1_ (t_1_-t_3_-strong response decliner) and ten a moderate response (t_1_-t_3_-moderate response decliner). The decrease of antibody response between t_1_ and t_3_ (5.7 months) and between t_2_ and t_3_ (2.8 months) is summarized in **supplemental table 2**. Taking into account the t_1_-t_3_ and t_2_-t_3_ time overlap, in total, 23 individuals have declined antibody responses between measurements at t_1_/t_2_ and t_3_ during a median time of 5.0 months. Overall, the RBD-and NP-specific antibody response of these 23 decliner has decreased by 19% per month for both. The monthly decline of antibody response was significantly correlated with the strength of response measured at t_1_/t_2_ with an r of 0.706 (p<0.001) for RBD and an r of 0.887 (p<0.001) for NP (**supplemental figure 1)**. Strong responders had a more pronounced monthly decline than moderate responder and the proportional decline between t_2_ and t_3_ was comparable to the one between t_1_ and t_3_ in spite of the shorter time span (**supplemental table 2**). Taking into account that exponential decline, the median half-life of RBD- and NP-specific responses were 5.5 [2.3-15.8] and 5.7 [2.2-11.2] months. In addition, the median time in which a positive antibody response (5 U/mL cut-off) for either RBD or NP can be maintained was 6.0 [1.6-19.8] months for all decliners and 10.2 [6.3-23.4] months for the strong-response decliner.

**Figure 3.**
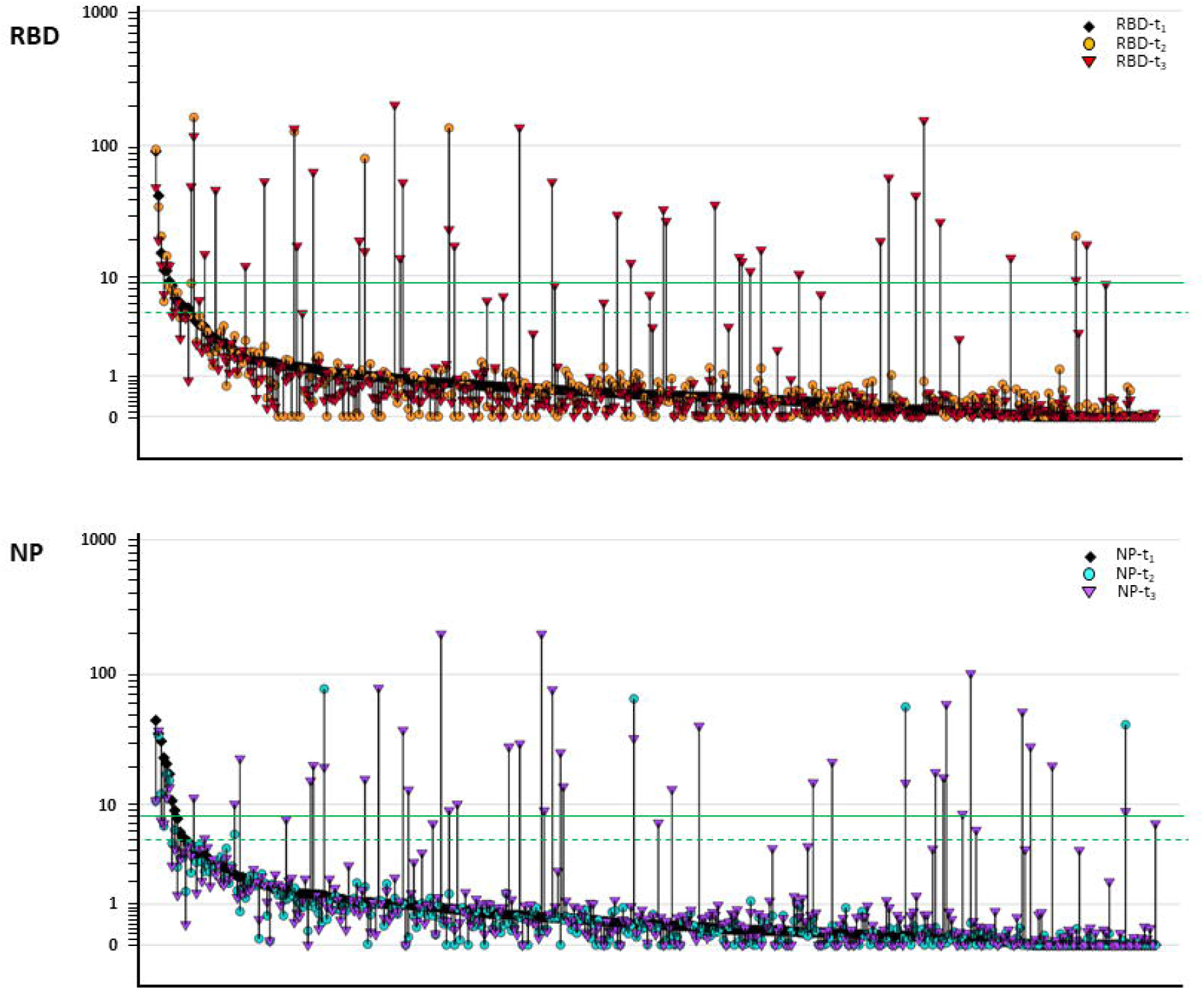
Shift of RBD- and NP-specific IgG response during study. SARS-CoV-2-specific IgG responses of study participants at time point t_1_ (black rhombs), are depicted ordered from high to low/background. The reference range (<5 U/mL) representing no response is separated from a moderate positive response (≥5 and <9 for anti-RBD and ≥5 and <8 for anti-NP) by a dashed green line and from a strong positive response (≥ 9 U/mL for anti-RBD and ≥ 8 U/mL for anti-NP) by a solid green line. The matching responses at t_2_ (circles), and t_3_, (triangles) are connected by a vertical line. RBD-specific responses are represented by orange (for t_2_) and red (for t_3_) symbols, NP-specific responses by turquois (for t_2_) and purple (for t_3_) symbols.

Of note, we did not find any elimination of a strong response between t_1_ and t_2_ or between t_1_ and t_3_. In contrast, out of the mentioned 12 moderate responders at t_1_ only 3 had still a moderate response at t_3_, 1 resigned, 1 converted to a strong response, and 7 did not reach the cut-off for moderate response at t_3_.

### Association of antibody response with RT-PCR data

Out of 395 HCW tested for SARS-CoV-2-specific IgGs, 249 have also been tested at least once for the presence of an acute infection with SARS-CoV-2 during the study by RT-PCR and 53 of these were positive. As mentioned above, applying ELISA, 73 out of all 395 HCW have been tested at least once positive for SARS-CoV-2-specific IgGs during the study. Thus, the number of HCW with ELISA-assessed positive antibody response is 38% higher (n=20) than infections detected by RT-PCR in the whole study population.

Taking into account only HCW who have been tested by both methods, RT-PCR and ELISA, we found that only four RT-PCR-positive HCW had no antibody response, reflecting an antibody response rate of 92% in RT-PCR-positive tested HCW. In contrast, only 73% of HCW with an antibody response have also been tested RT-PCR-positive (46/63). Regarding a strong antibody response, only 83% have been tested RT-PCR-positive (43/52).

### Association of antibody response with COVID-19-symptoms and further parameters

Taking into account the survey data, HCW who had COVID-19-symptoms at t_3_ were significantly more likely to be seropositive than asymptomatic ones (36% vs. 8% p<0.001), but this was not the case at t_1_ (p=0.193) or t_2_ (p=0.645). Further, there was no significant difference between male and female HCW being seropositive at any time point (21% vs. 18%, p=0.518) or between HCW with a BMI ≥25 compared to those with BMI <25 (22% vs. 17%, p=0.226). HCW above 40 years had a similar prevalence compared to younger (≤40 years) ones (16% vs. 18%, p=0.603) and participants sharing their household with children or adolescents younger than 25 years had no significantly increased risk for being seropositive compared to participants without younger persons in their households (19% vs. 14%, p=0.202). HCW working at a regular hospital had a slightly but not significantly lower prevalence than those at a COVID-19-specialized hospital (14% vs. 21%, p=0.068) and also smokers had a lower prevalence, which just failed significance (9% vs. 18%, p=0.060).

## Discussion

### Main findings

In our study the antibody response was clearly higher after the second massive wave compared to the first wave reflecting the incidence rate in Austria (**figure 1** and (15)). Of note, the number of undetected SARS-CoV-2 infections during our study was quite high as only 83% of HCW with a strong antibody response, have previously been identified by RT-PCR.

Moreover, a conversion to a strong response during the study was much more likely than conversion to a moderate response only and a strong response was more stable than a moderate response.

A further important finding was that we found no elimination of a strong response during the study: All participants with a strong response maintained a positive response during the study and, according to extrapolation, will keep it for 10 months. Similarly, the half-life of positive antibody responses was about six months for both, the RBD- and NP-specific response.

### Seroprevalence after the first wave in the light of other study data on HCW

Our data revealing a 3% seroprevalence at t_1_, after the first wave, are slightly above those from HCW in Germany (20, 21) and Italy, apart from the North (22, 23) being in the range of 1–2% around the same time. Higher rates of 5-6% were seen in the Veneto Region, Italy (24), Belgium (25), Norway (26) and Northern England, UK (27). One of the highest incidence rates of COVID-19 infections in the world were seen in the U.S., with and a seroprevalence rate of 19% in the general population (28) and 27% in HCW at the New York City Hospital South Bronx at the same time (29). Almost similar rates were found in HCW in Sweden (19%) (30) and some parts of the U.K. namely London (32%) (31) and Birmingham (24%) (32). Nevertheless, these rates are still below the seropositivity rate of 67%, which has initially been estimated as threshold for community immunity against SARS-CoV-2 (33) and now has been estimated to be as high as 85% according to CDC (34).

### Seroprevalence at the beginning and at the end of the second wave

A recent seroprevalence study of the general population in Austria comprising 2229 participants and collecting samples between 12^th^ to 14^th^ November, which took place during the second wave found neutralizing antibodies in 92 samples reflecting a seroprevalence of 4.7% (35). This is just matching our data about the same time (t_2_) and thus proposes that HCW in Vorarlberg were well prepared facing the challenges by COVID-19 in the local health care system although they might have a higher chance of being infected than the general population. Passing the second wave, Austria had one of the highest incidence rates in the world (4) and the seroprevalence after the second wave has been hypothesized to be about 15% in the general population (36). Around the same time, at t_3_ of our study, we found a massive increase to 14 %, having a strong antibody response. This proposes again that HCW in Vorarlberg may have had an infection rate comparable to the general population. As all HCW in Vorarlberg had the opportunity for vaccination starting at 7^th^ January, it remains speculative whether the seroprevalence might have further increased or plateaued.

### Seroconversion, protection and reinfection

Even though our study primarily aimed at observing the prevalence of seroconversion of all HCW during first and second wave of the COVID-19 pandemic, focusing only on the subgroup of responder we found that, a strong response was more stable than a moderate response.

These findings are in good alignment with the very fast increase in antibody titers and neutralization within only 10 days after symptom onset, tested with the same assay (19). All participants who once have developed a strong response maintained a positive response, either still a strong one or at least a moderate one, during the full study time. An extrapolation thus suggests that these strong responders will keep their response for about ten months.

This is in line with previous data of recent studies in the UK and Spain, demonstrating that SARS-CoV-2 infection-acquired immunity is present for at least six months (14, 25) and suggesting that protective immunity will last up to a few years (14). A further study in New York City has found only a moderate decline regarding the spike protein-specific response during five months (10). We here report a mean decline of 51% and 60% during five months for RBD- and NP-specific responses, respectively. A decrease of 17 % and 31 % for anti-spike IgG and anti-NP IgG titers has been reported in a study comprising 847 workers at Institute Curie in Paris, France during 4-8 weeks thus accounting a rather short-lived immune responses with only 87 days for anti-spike IgG and 35 days for anti-NP IgGs, respectively (12). Wajnberg et al. have suggested that the stability of the antibody response over time may depend on the serologic target (10) with a faster decline of NP compared to RBD. Other than NP, the spike protein is the main and potentially the only target for neutralizing antibodies (37). It thus appears that the RBD is more suited than NP for surveillance of long-term immune response by ELISA. Nevertheless, RBD-specific IgG response as investigated in our study as well as in most others on seroprevalence is only a fragment of the very complex post-infection immunity and longevity of response.

Finally, we also have noticed one case in which a moderate antibody response at t_1_ has converted to a strong response at t_3_, most likely representing a reinfection. That said, the number of responders at t_1_ and t_2_ is small compared to the initial study number and thus the conclusions (including those regarding reinfection, immunity, elimination time, and half-life) for this subgroup are limited and should be taken with care. Further limitations are mentioned in the following.

### Limitations

This study is not a random sample of either the general population or the HCW of Vorarlberg. Included are voluntary and of age HCW. Results are not necessarily applicable to the general population and especially not to children or elderly people (≥ 65 years). At least some results should be interpreted with caution, as it is possible that some of our participants which have been classified to “no response” due to a response below the assay cut-off of <5 U/mL were infected with SARS-CoV-2 a few months before sampling, and either have had only a low antibody response post-infection, and/or have already dropped below the assay threshold. Apart from that, our study only gives information about post-infection antibody-response and not about immunity or the chance of reinfections. In that context, it is impossible to fully explain the nature of change of antibody-specific responses in our study, e.g. for responders of which some may be impacted by a secondary contact to the virus thus acting as kind of a booster. Furthermore, it has already been demonstrated that a NP- or spike-specific antibody response may not always be present following a proven SARS-CoV-2 infection (12). Apart from that, a large variety of different commercial ELISAs has been used for the above-mentioned serological study data. Although IgG-specific ELISAs have been proposed to be appropriate for prevalence testing, accuracy significantly differs between different serological testing methods (38), thus, the between-comparison should always be done with caution. Finally some participants have been vaccinated during sampling at t_3_, but in no case vaccination took place more than one week before sampling. Antibody responses are generally not mounted within one week after vaccination (39), and converters at t_3_ who have been vaccinated had responses for RBD and NP thus we preclude an effect of the RBD-based vaccine.

### Implications for health policy and clinical perspectives

Given the limitations mentioned above, the antibody response is yet widely used as a surrogate for deciding whether post-infection immunity to SARS-CoV-2 exists and the antibody response in our study has proven to persist for several months. That said, our and others’ findings do not support exempting those positive for anti-SARS-CoV-2 antibodies from current infection control, other public health constraints, or the ongoing vaccination. Anyway, the current seroprevalence of HCW is far beyond any herd immunity threshold Our findings suggest serological testing as routine application for determining and monitoring the success of detecting acute infections and is therefore an important tool managing the ongoing COVID-19 pandemic. Given the 38% higher number of HCW with antibody response than RT-PCR-verified infections assessed by routine surveillance, and the at least 17% undetected infections of HCW in our hospitals recommends a massively increased infection surveillance, either by routine serological, PCR-based, other test strategies e.g. daily lateral flow tests. Apart from that, further studies are necessary to determine the long-time duration of post-infection antibody response and immunity and compare it to vaccination data as this has major implications for the future of the current SARS-CoV-2 pandemic and public health system. For the particular study participants, the ELISA may also be very helpful for determining the success of vaccination. We plan to continue this longitudinal serosurvey for several months analyzing the antibody response after vaccination started in Austria at January 2021.

## Data Availability

The data that support the findings of this study are available from the corresponding author upon reasonable request.

## Acknowledgments

We are grateful to the Vorarlberger Landesregierung (Bregenz, Austria) for continuously supporting our research institute. We are also grateful to all state hospitals in Vorarlberg and in particular to the Institute of Pathology at the Academic Teaching Hospital Feldkirch for their support. The present study was funded by the Austrian Research Promotion Agency (FFG).

