## Supplemental data for "SARS-CoV-2 serostatus of healthcare worker in the Austrian state Vorarlberg between June 2020 and January 2021"

**Supplemental material**

**Supplemental table 1**

**Residence and profession**

| **Residence** | Vorarlberg | 364 (92.2%) |
| --- | --- | --- |
|  | out of Vorarlberg | 14 (3.5%) |
|  | not specified | 17 (4.3%) |
|  | total | 395 (100%) |
| **Country of Birth** | Austria | 300 (75.9%) |
|  | Germany | 38 (9.6%) |
|  | Italy | 12 (3.0%) |
|  | Other EU | 11 (2.8%) |
|  | Outside EU | 10 (2.5%) |
|  | not specified | 24 (6.1%) |
|  | total | 395 (100%) |
| **Professional role** | Reception | 10 (2.5%) |
|  | Secretarial | 18 (4.6%) |
|  | Physician | 96 (24.3%) |
|  | Nursing/Physio | 250 (63.3%) |
|  | Radiology | 10 (2.5%) |
|  | Service | 9 (2.3%) |
|  | Lab | 1 (0.3%) |
|  | not specified | 1 (0.3%) |
|  | total | 395 (100%) |

**Supplemental table 2**

**Seroconversion and decline of antibody response during study**

|  |  | **Change of response** | **Change of response per month** | **Half-life in months** |
| --- | --- | --- | --- | --- |
| **t_1_-t_3_ all HCW**  **(n=371)** | RBD  NP | +4.0 U/mL (335 %)  +3.4 U/mL (270 %) | n.a.  n.a. | n.a.  n.a. |
| **t_1_-t_3_-strong response converter (n=44)** | RBD  NP | +35.9 U/mL (4233 %)  +29.8 U/mL (4368 %) | n.a.  n.a. | n.a.  n.a. |
| **t_1_-t_3_-moderate response converter (n=6)** | RBD  NP | +4.0 U/mL (349 %)  +2.6 U/mL (231 %) | n.a.  n.a. | n.a.  n.a. |
| **all t_1_-t_3_-converters**  **(n=50)** | RBD  NP | +32.1 U/mL (3634 %)  +26.5 U/mL (3611 %) | n.a.  n.a. | n.a.  n.a. |
| **t_1_-t_3_ strong response-decliner (n=9)** | RBD  NP | - 7.8 U/ml (- 38 %)  - 11.7 U/ml (- 52 %) | - 1.5 U/mL (- 7 %)  - 2.1 U/mL (- 9 %) | 7.5 [4.5-215.4]  3.4 [2.7-11.5] |
| **t_1_-t_3_ moderate response-decliner (n=10)** | RBD  NP | - 1.5 U/ml (-38 %)  - 1.1 U/ml (- 36 %) | - 0.3 U/mL (- 7 %)  - 0.2 U/mL (- 6 %) | 5.6 [2.0-17.2]  7.6 [6.1-40.9] |
| **all t_1_-t_3_-decliner**  **(n=19)** | RBD  NP | - 4.5 U/mL (- 38 %)  - 6.1 U/mL (- 50 %) | - 0.8 U/mL (- 7 %)  - 1.1 U/mL (- 9 %) | 5.7 [3.8-17.2]  6.2 [2.9-17.3] |
| **t_2_-t_3_ strong response-decliner (n=11)** | RBD  NP | - 27.8 U/ml (- 54 %)  - 16.3 U/ml (- 53 %) | - 11.9 U/mL (- 23 %)  - 6.7 U/mL (- 21 %) | 2.9 [0.9-4.6]  4.0 [1.5-17.6] |
| **t_2_-t_3_ moderate response-decliner (n=7)** | RBD  NP | - 1.1 U/ml (-23 %)  - 0.4 U/ml (- 18 %) | - 0.4 U/mL (- 7 %)  - 0.1 U/mL (- 6 %) | 11.0 [1.4-127.6]  10.6 [5.3-41.3] |
| **all t_2_-t_3_-decliner**  **(n=18)** | RBD  NP | - 17.5 U/ml (- 52 %)  - 10.1 U/ml (- 51 %) | - 7.4 U/ml (- 22 %)  - 4.1 U/ml (- 21 %) | 3.5 [1.4-11.5]  5.1 [2.5-31.0] |
| **all strong response decliner**  **(n=13)** | RBD  NP | - 23.3 U/mL (- 52 %)  - 20.9 U/mL (- 61 %) | - 9.0 U/mL (- 20 %)  - 6.7 U/mL (- 20 %) | 5.3 [1.8-14.5]  2.7 [1.8-5.1] |
| **all moderate response decliner (n=10)** | RBD  NP | - 1.5 U/mL (- 38 %)  - 1.1 U/mL (- 36 %) | - 0.3 U/mL (- 7 %)  - 0.2 U/mL (- 6 %) | 5.6 [2.0-17.2]  7.6 [6.1-40.9] |
| **all decliner**  **(n=23)** | RBD  NP | - 13.8 U/mL (- 51 %)  - 12.3 U/mL (- 60 %) | - 5.2 U/mL (- 19 %)  - 3.9 U/mL (- 19 %) | 5.5 [2.3-15.8]  5.7 [2.2-11.2] |

The table summarizes decline as well as raise of antibody response for the respective time interval. Converters had an increase of antibody response from background to either moderate or strong. Decliners were defined as not converters and having either a decrease of a strong or moderate antibody response or no change of a strong or moderate antibody response. Median half-lives, given with interquartile range, were calculated assuming an exponential decline if applicable and are given in month until half of the initial response is lost.

**Supplemental figure 1**

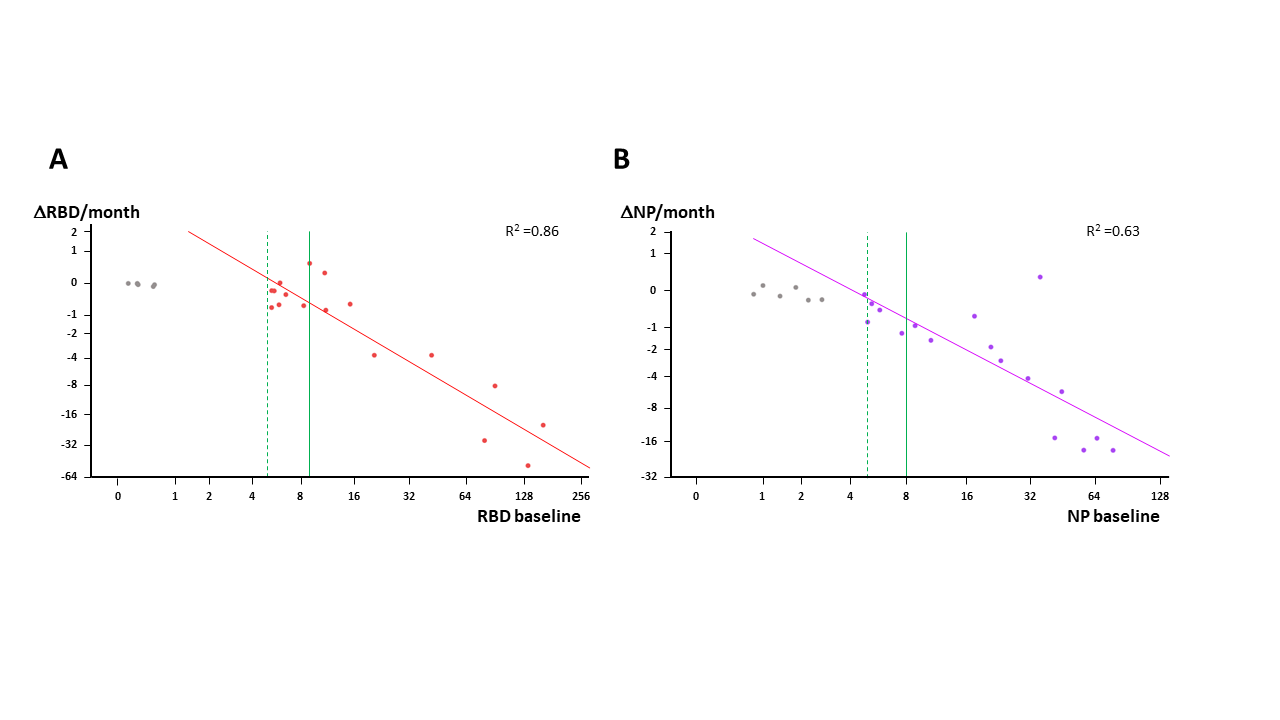

**Supplemental figure 1: Monthly decline of IgG response in correlation with baseline IgG response**

The monthly decline of the SARS-CoV-2-specific response of study participants in relation to their response at baseline is depicted for anti-RBD-specific (A) and for anti-NP-specific IgGs (B). A reference range of 0-5 U/mL representing no response is separated from a moderate positive response (≥5 and <9 for anti-RBD and ≥5 and <8 for anti-NP) by a dashed green line and from a strong positive response (≥ 9 U/mL for anti-RBD and ≥ 8 U/mL for anti-NP) by a solid green line. Grey dots represent values outside the positive range and were excluded for calculation of the regression lines given as solid red and turquois lines with R^2^ indicated.
